# ST-segment depression on resting electrocardiogram in patients with out prior cardiac disease — a comparison with stress electrocardiogram and echocardiography —

**DOI:** 10.1101/2023.05.03.23289480

**Authors:** Masaki Morita

## Abstract

**Background:** The clinical significance of ST-segment depression (STD) on electrocardiogram (ECG) in resting patients without prior cardiac disease and symptoms of angina pectoris in unclear. This study was aimed to elucidate this problem.

**Methods:** Patients with non-anginal chest pain or those who underwent ECG screening were enrolled. Echocardiography (Echo) and resting and stress ECG were performed.; stress ECG was recorded during exercise and recovery period. For stress ECG, ST-segment/heart-rate loop was adopted. The clinical results in patients with and without STD on resting ECG were compared.

**Results:** Eighty-two patients were included; a mean age of 61.4 ± 14.1 years old. ECG revealed that 13 (15.9%) patients had left ventricular hypertrophy (LVH; Sokolow-Lyon’s voltage > 3.5 mV), whereas 34 (41.5%) patients had STD (ST-segment voltage at the J point < –0.05 mV). Echo revealed a mean left ventricular mass (LVM) of 182 ± 74 g. The stress ECG revealed the mean recovery index was –1.2° ± 19.1°. A correlation of STD voltage with Romhilt-Estes score on resting ECG (r = –0.34, p = 0.0015) and LVM on Echo (r = –0.26, p = 0.018) were observed. From ROC curves, STD tended to diagnose more accurately than other ECG parameters. From multiple logistic analysis, age, Romhilt-Estes score and peak STD on stress ECG were independently correlated with STD on resting ECG.

**Conclusion:** In the patients without prior cardiac disease and anginal chest pain, STD on resting ECG was suggestive of older age, LVH on ECG (independent) and LVH on Echo (not independent).

**Registration:** URL: https://www.umin.ac.jp/ctr/index.htm; Unique identifier: UMIN000050291

**WHAT IS KNOWN:** ST-segment depression (STD) on resting electrocardiogram (ECG) among the subjects without prior cardiac disease and symptoms of angina is suggested to imply left ventricular hypertrophy.

**WHAT THE STUDY ADDS:** In order to investigate the significance of STD on resting ECG among the subjects without history of cardiac disease or anginal chest pain, especially to rule out the incidence of silent myocardial ischemia, echocardiogram and stress ECG were contemporarily performed for them.

## 1. Introduction

Electrocardiogram (ECG) is one of the most frequently used diagnostic techniques in clinical practice. It is useful for detecting myocardial ischemia, conduction disturbances, and arrhythmias. It can also be used to examine various types of treatments, assess cardiovascular risk for intermediate and high-risk surgeries, and evaluate individuals in high-risk occupancies and athletes^1^.

ST-segment and T-wave changes are one of the most common ECG findings^2^ and can reveal the ischemic and non-ischemic origins of a cardiac event^3^. ST-segment and T-wave changes are also observed in patients without history of prior cardiovascular disease or any symptoms of angina. Interpretation of these changes is challenging, and given their low specificity, they are called “non-specific ST-T changes^2^.”

However, ST-segment depression (STD) and T-wave inversion on resting ECG could be indicative of the risk for postoperative myocardial ischemia or even death^4, 5^. Furthermore, they could also predict future cardiovascular events or diseases^6, 7^.

Therefore, patients with ST-segment and T-wave changes should be carefully examined. The presence of left ventricular hypertrophy (LVH)^8^ on echocardiography (Echo) and silent myocardial ischemia on stress ECG should be assessed in routine practice.

Concurrent use of exercise stress ECG and Echo with resting ECG can identify cardiac diseases based on changes in ECG waveforms. However, very few studies have compared these three methods, especially among the patients who show ST-T changes on resting ECG, without symptoms of or history of prior cardiac diseases.

This study aimed to evaluate the significance of STD on resting ECG with no history of cardiac disease by combining contemporary stress ECG testing and Echo.

## 2. Materials and Methods

### 2.1 Study Population

This study was retrospective, observational study for the patients who were consecutively enrolled in the following inclusion criteria. Eighty-two patients without a history of prior cardiac disease were enrolled in this study who attended the author’s clinic.

Patients presenting with following were included; non-anginal chest pain, presence of STD, screening of covert cardiac disease due to high-risk non-cardiac surgery and colonoscopy.

Exclusion criteria were: clinical signs of acute coronary syndrome, typical angina pectoris; syncope; congestive heart failure; ECG showing left or right bundle branch blocks; pre-excitation syndrome; pacemaker implantation; any degree of atrioventricular blocks, and tachyarrhythmias (paroxysmal supraventricular tachycardia, sustained or non-sustained ventricular tachycardias). However, four patients with atrial fibrillation without a history of congestive heart failure or ischemic heart disease were enrolled in this study. All patients underwent ECG at rest, stress ECG using cycle ergometer stress test or treadmill test and Echo to assess non-anginal chest symptoms for cardiac diseases.

### 2.2 Resting Electrocardiogram

ECG was performed in all patients at rest in the supine position, in a separated and calm examination room by a laboratory technician who was blinded to the other data. The paper speed was 25 mm/s, and calibration of the electrical amplitude was 10 mm, which was equal to 1 millivolt (mV). In each ECG, the height of the R wave on the V_5_ lead and the depth of the S wave on the V_1_ lead were calculated to measure the Sokolow-Lyon’s voltage, and the voltage of STD and T-wave inversion were determined as the one observed non the deepest leads. Heart rhythm was categorized into normal sinus rhythm or atrial fibrillation. The criteria for the presence of LVH and STD were as follows; Sokolow-Lyon’s voltage > 3.5 mV, voltage of STD at the J point < –0.05 mV. The Romhilt-Estes score was also determined using ECG, as measured and verified by the author.

### 2.3 Echocardiography

Echo was performed by a sonographer blinded to the other data. The Echo was examined with the patients in the supine and right anterior oblique positions. Longitudinal, left parasternal view, interventricular septal wall thickness at end-diastole, left ventricular posterior wall thickness at end-diastole (LVPWTd) and left ventricular internal diameter at end-diastole were determined. Left ventricular mass (LVM) was calculated using the following formula^9^

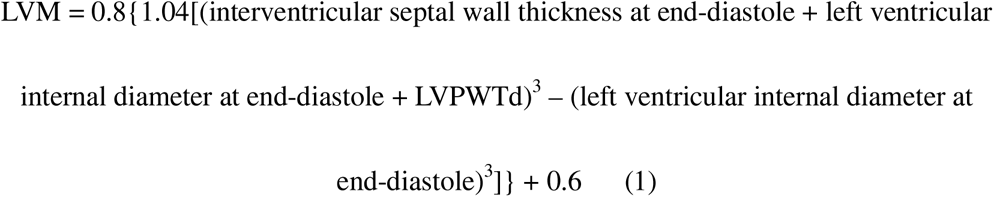

LVH on Echo was considered in case of the following findings; LVM > 163 g in women or > 225 g in men^9^. Doppler recording was also obtained to evaluate the significant acceleration of transvalvular flow and transvalvular regurgitant flow.

### 2.4 Exercise stress electrocardiogram

Exercise stress test was conducted on all patients in another separated and calm examination room by another laboratory technician who was blinded to the other data. Seventy-two patients underwent cycle ergometer test and 10 patients underwent treadmill test. Symptom-limited, incremental workload protocols were adopted for stress ECG. On cycle ergometer test, workload increased 15∼50 W every 2 minutes, according to the patient’s age and/or physical fitness. On treadmill test, Sheffield protocol or Bruce protocol was used according to the patient’s age or fitness.

Blood pressure, heart rate and 12 lead ECG were examined before, during and after stress test and described every one minute.

On the stress ECG, an analysis of ST-segment/heart-rate loop method was used to achieve a more accurate diagnosis of coronary artery disease (CAD)^10, 11^. On the graphic chart of ST-segment/heart-rate loop, the slope of the line connecting the STD threshold to maximal STD during exercise (line 1) was defined as ST-segment/heart-rate slope (ST/HR slope)^12^.

Subsequently, as a method for determining heart-rate -adjusted STD changes during the recovery period, the recovery index was defined as the angle between the line 1 and the line connecting the maximal STD during exercise to a point of 30 s into recovery (line 2) (Figure 1)^13^. The results were considered positive (ischemic) with ST/HR slope > 3.5 μV/beats per minute and recovery index > 0°.

**Figure 1.**
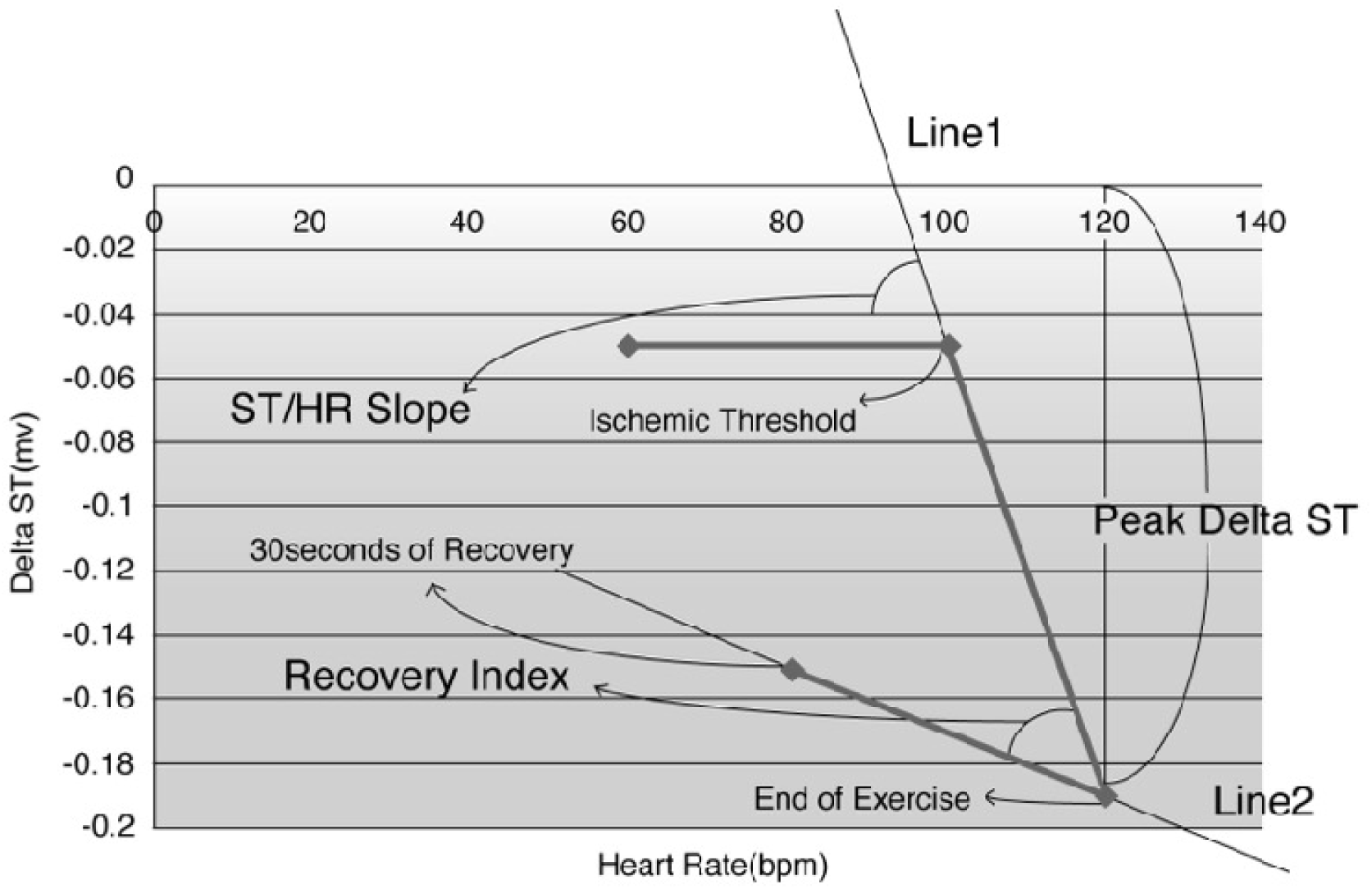
Pattern diagram of ST-segment/heart-rate loop analysis for assessing exercise stress electrocardiographic findings. Abbreviations: bpm, beats per minute; Delta ST, voltage of ST-segment depression; mv, millivolt

### 2.5 Statistical Analysis

Values were reported as mean ± standard deviation for numerical data and percentages for categorical data. The Welch’s test or Pearson’s chi-square test was used to compare patients, and linear regression analysis were made for evaluating the correlation. Receiver operating characteristic (ROC) curves were used to determine the optimal parameters for diagnosis. Multivariate logistic regression analysis was made to test the independence of the parameters, which revealed a correlation in univariate analysis. The statistical software BellCurve for Excel (Social Survey Research Information, Co. Ltd, Tokyo, Japan) was used for statistical analysis. Statistical significance was set at p < 0.05.

## 3. Results

### 3.1 Clinical characteristics

A total of 83 patients were recruited for this study. Of the patients, 45% were in their sixties, 27% were in their seventies. Three-quarters of patients had non-anginal chest pain or were asymptomatic (Figure 2). Moreover, ECG evaluation revealed that one-sixth of the patients had LVH, and two-fifths had STD < –0.05 mV. The mean Sokolow-Lyon’s and STD voltages were within normal ranges. The Shapiro-Wilk test was used to determine STD voltage normality; it was not normally distributed (W = 0.87, p < 0.001), so a non-parametric test was used to compare data between the patients who had STD and normal ST-segment (Table 1).

**Figure 2.**
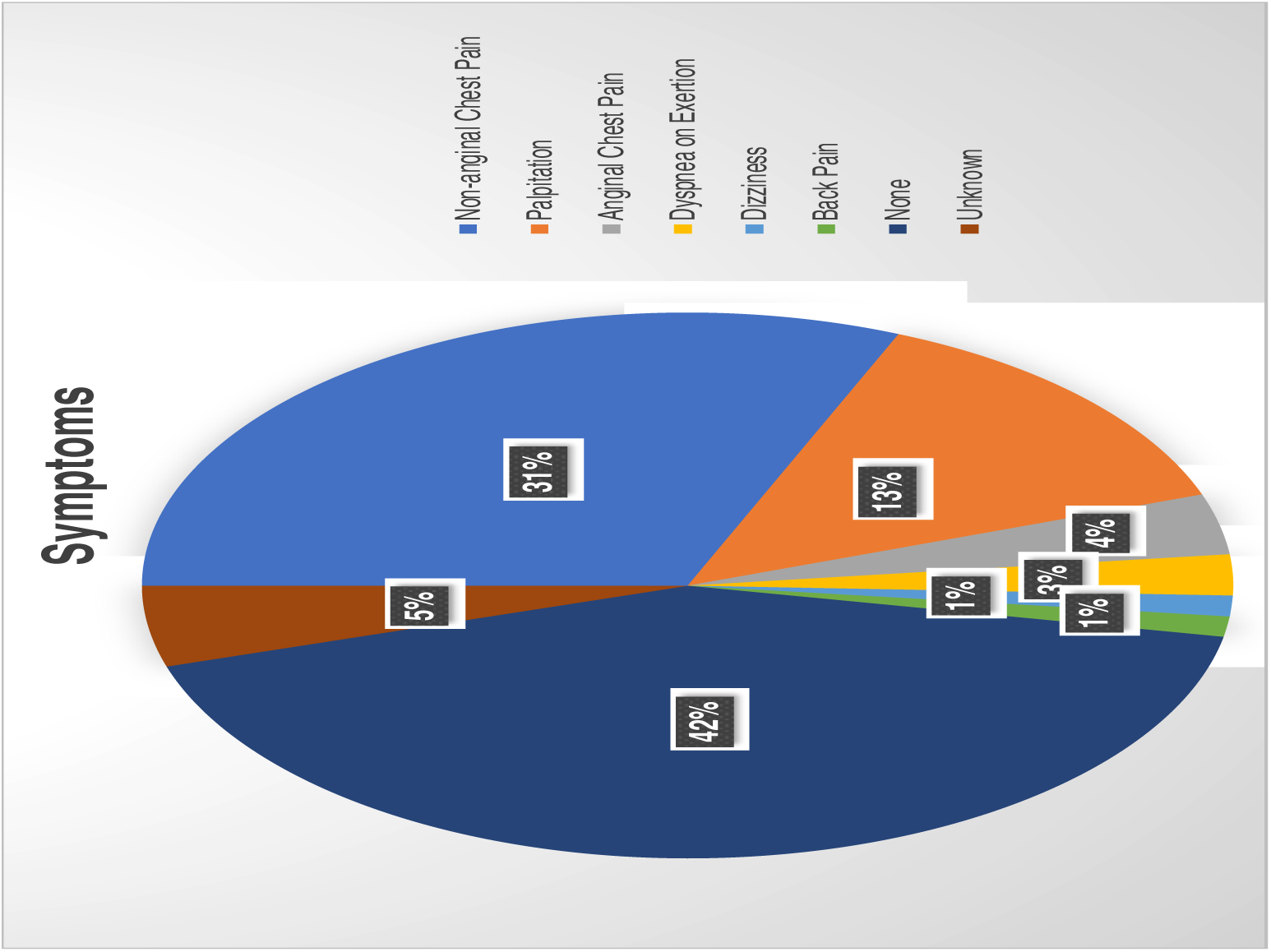
Distribution of patient’s

**Table 1.** Baseline characteristics and resting electrocardiographic parameters: In the entire group, and normal ST-segment and ST-segment depression groups.

| Parameters | Entire (n =82) | STN (n =48) | STD (n = 34) | p-value |
| --- | --- | --- | --- | --- |
| Age | 61.4 ± 14.1 | 59.4 ± 16.6 | 64.8 ± 8.4 | 0.17 |
| Gender (Men) | 36 (43.9%) | 23 (47.9%) | 13 (38.2%) | 0.10 |
| Hypertension | 42 (51.2%) | 22 (47.8%) | 20 (58.8%) | 0.06 |
| Diabetes mellitus | 14 (17.1%) | 6 (12.0%) | 8 (23.5%) | 0.06 |
| Dyslipidemia | 23 (28.0%) | 13 (27.1%) | 10 (29.4%) | 0.98 |
| Rhythm (SR) | 78 (95.1%) | 46 (95.8%) | 32 (94.1%) | 0.62 |
| Heart rate (bpm) | 72.2 ± 15.0 | 71.4 ± 14.0 | 73.3 ± 16.5 | 0.66 |
| ECG LVH | 13 (15.9%) | 7 (14.6%) | 6 (17.6%) | 0.60 |
| Sokolow-Lyon's<br>voltage (mV) | 2.8 ± 1.0 | 2.8 ± 1.1 | 2.9 ± 0.9 | 0.45 |
| Romhil-Estes score | 2.3 ± 2.0 | 1.9 ± 1.6 | 2.8 ± 2.3 | 0.086 |
| Voltage of STD (mV) | -0.037 ± 0.036 | -0.014 ± 0.016 | -0.072 ± 0.028 | < 0.001 |
| T wave change<br>(Inversion) | 3 (3.8%) | 0 (0%) | 3 (8.8%) | - |
| Note: p-values are obtained by comparison of normal ST-segment and ST-segment depression by Welch test or Pearson's chi-square test. Abbreviations: bpm, beats per minute; ECG LVH, electrocardiographic left ventricular hypertrophy determined by Sokolow-Lyon's voltage > 3.5 mV; SR, sinus rhythm; STD, ST-segment depression; STN, normal ST-segment |  |  |  |  |

Echo analysis revealed that the mean LVPWTd and mean LVM were in normal ranges. On stress ECG, the mean ST/HR slope and recovery index were in normal ranges (Table 2).

**Table 2.**
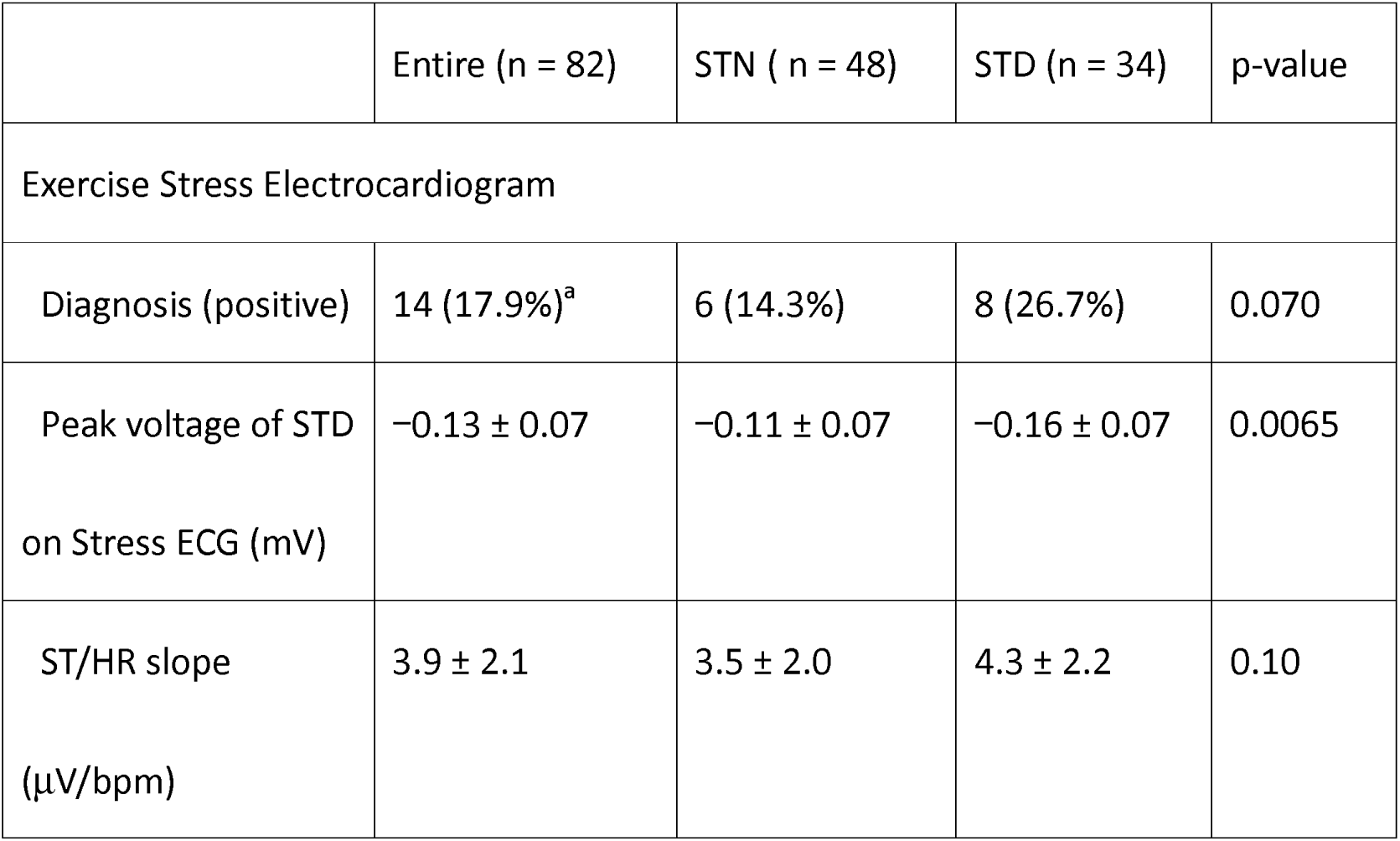
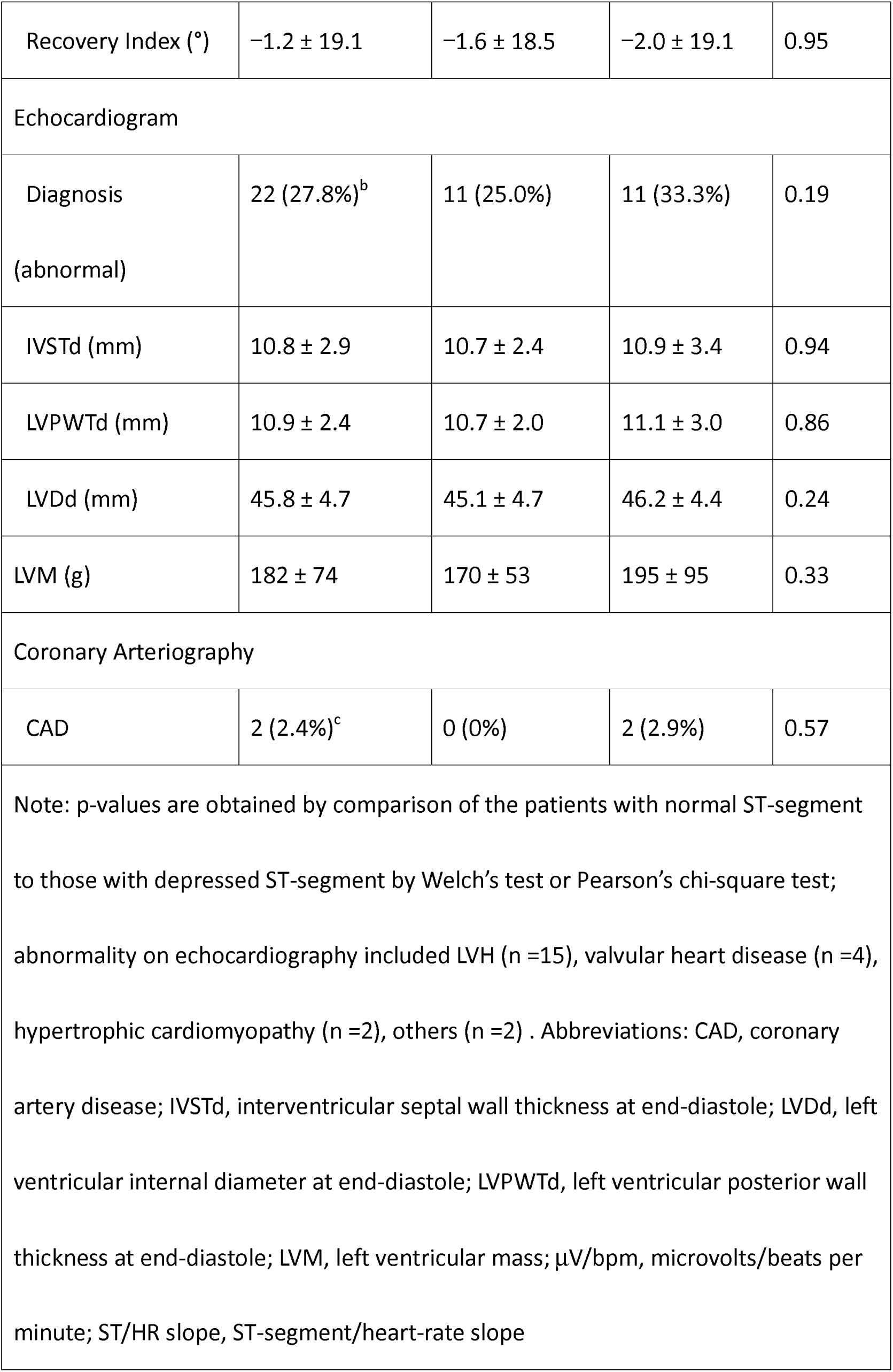
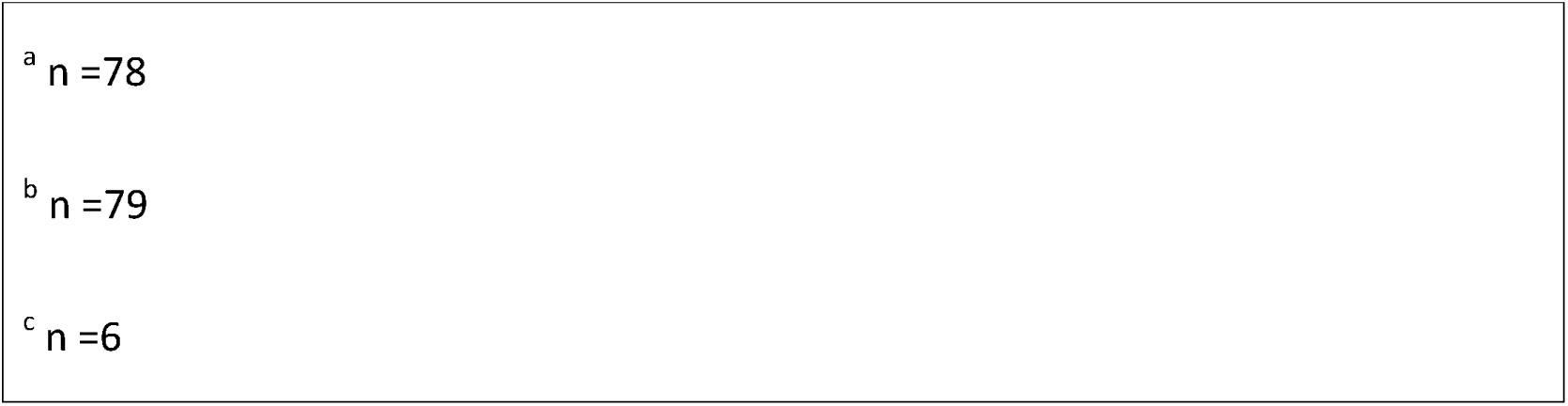
Cycle ergometer stress electrocardiographic, echocardiographic and coronary arteriographic parameters: In the entire group, and normal ST-segment and ST-segment depression groups.

|  | Entire (n = 82) | STN ( n = 48) | STD (n = 34) | p-value |
| --- | --- | --- | --- | --- |
| Exercise Stress Electrocardiogram |  |  |  |  |
| Diagnosis (positive) | 14 (17.9%)<sup>a</sup> | 6 (14.3%) | 8 (26.7%) | 0.070 |
| Peak voltage of STD<br>on Stress ECG (mV) | $-0.13 \pm 0.07$ | $-0.11 \pm 0.07$ | $-0.16 \pm 0.07$ | 0.0065 |
| ST/HR slope<br>(μV/bpm) | $3.9 \pm 2.1$ | $3.5 \pm 2.0$ | $4.3 \pm 2.2$ | 0.10 |
| Recovery Index (°) | -1.2 ± 19.1 | -1.6 ± 18.5 | -2.0 ± 19.1 | 0.95 |
| Echocardiogram |  |  |  |  |
| Diagnosis<br>(abnormal) | 22 (27.8%)<sup>b</sup> | 11 (25.0%) | 11 (33.3%) | 0.19 |
| IVSTd (mm) | 10.8 ± 2.9 | 10.7 ± 2.4 | 10.9 ± 3.4 | 0.94 |
| LVPWTd (mm) | 10.9 ± 2.4 | 10.7 ± 2.0 | 11.1 ± 3.0 | 0.86 |
| LVDd (mm) | 45.8 ± 4.7 | 45.1 ± 4.7 | 46.2 ± 4.4 | 0.24 |
| LVM (g) | 182 ± 74 | 170 ± 53 | 195 ± 95 | 0.33 |
| Coronary Arteriography |  |  |  |  |
| CAD | 2 (2.4%)<sup>c</sup> | 0 (0%) | 2 (2.9%) | 0.57 |
| Note: p-values are obtained by comparison of the patients with normal ST-segment to those with depressed ST-segment by Welch's test or Pearson's chi-square test; abnormality on echocardiography included LVH (n =15), valvular heart disease (n =4), hypertrophic cardiomyopathy (n =2), others (n =2) . Abbreviations: CAD, coronary artery disease; IVSTd, interventricular septal wall thickness at end-diastole; LVDd, left ventricular internal diameter at end-diastole; LVPWTd, left ventricular posterior wall thickness at end-diastole; LVM, left ventricular mass; μV/bpm, microvolts/beats per minute; ST/HR slope, ST-segment/heart-rate slope |  |  |  |  |

<sup>a</sup> n =78
<sup>b</sup> n =79
<sup>c</sup> n =6

### 3.2 Locations of ST-segment depression on resting electrocardiogram

Locations of ST-segment changes are shown (Figure 3). Two-thirds of the patients had STD on both inferior and lateral leads, one-ninth had STD only on the inferior leads, one-fifth had STD only on lateral leads, and none had STD in the precordial leads.

**Figure 3.**
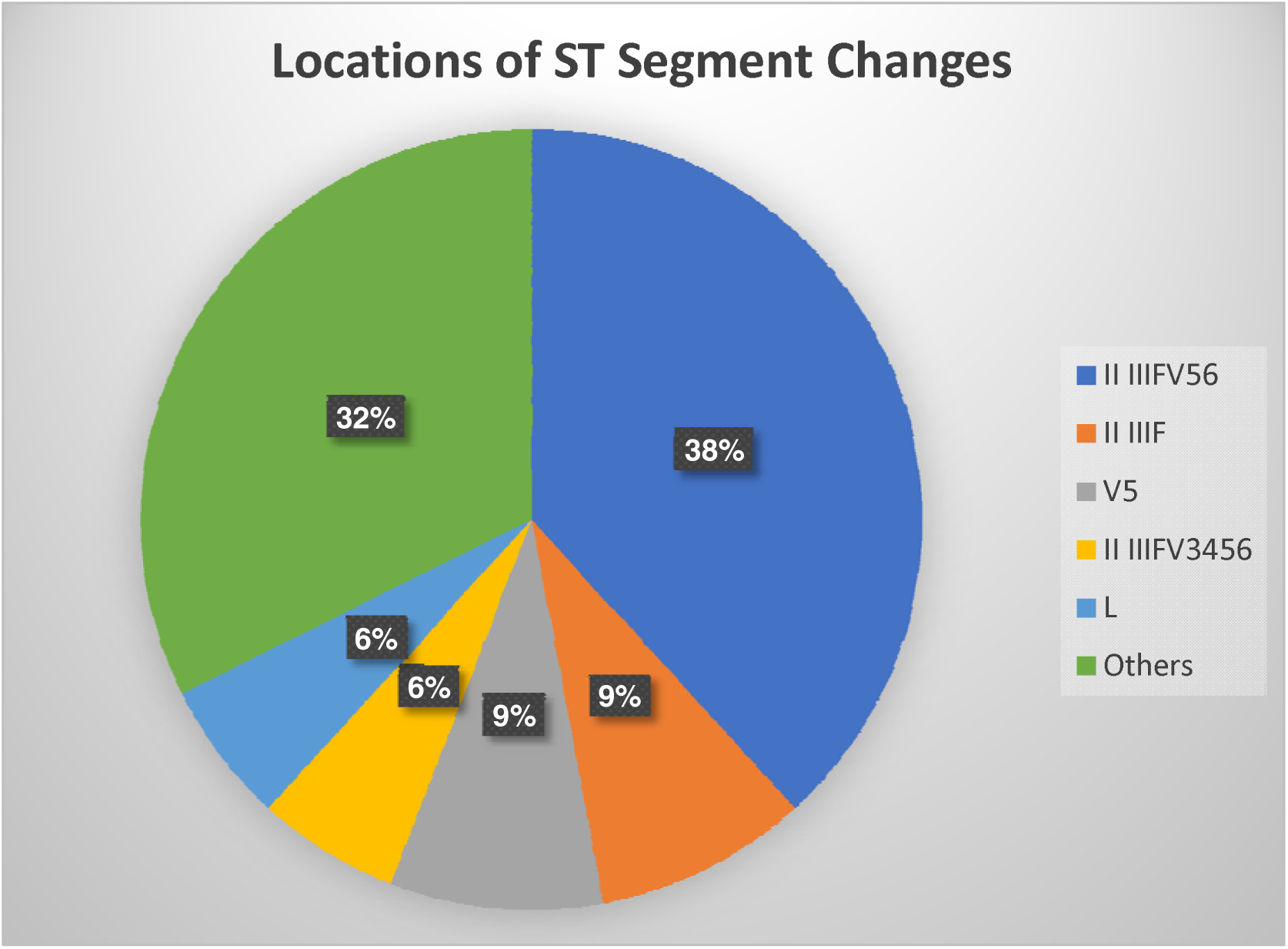
Locations of ST-segment changes

### 3.3 Comparison of electrocardiographic parameters between patients with normal and depressed ST-segments

The voltage of STD on resting and stress ECG were significantly different between STD and normal ST-segment patients; no differences were found between them for other parameters (Tables 1 and 2).

### 3.4 Correlation between the ST-segment depression voltage of resting electrocardiogram and parameters of resting and stress electrocardiogram and echocardiography

STD voltage on resting ECG correlated with the Romhilt-Estes score on ECG at rest, peak voltage of STD during cycle ergometer stress ECG. STD voltage was also associated with LVPWTd and LVM on Echo. However, it did not correlate with the ST/HR slope and recovery index of ECG on the stress ECG; ST/HR slope and recovery index of STD patients was smaller in proportion to the changes of STD during stress ECG test (Table 3). Therefore, the patients with STD had deeper ST-segment depression on stress ECG, but when considering heart-rate-adjusted ST-segment changes during exercise, the stress ECG of patients with resting STD showed negative (non-ischemic) STD (Tale 2) changes during and in the recovery period of exercise stress ECG.

**Table 3.** Correlation of voltage of ST-segment depression on resting electrocardiogram with parameters of basic characteristics, resting and exercise stress electrocardiogram and echocardiogram.

|  |  | r | p-value |
| --- | --- | --- | --- |
| Basic characteristics | Age | -0.25 | 0.024 |
|  | Heart rate | -0.17 | 0.13 |
| Resting ECG | Sokolow-Lyon's voltage | -0.046 | 0.68 |
|  | Romhilt-Estes score | -0.34 | 0.0015 |
| Exercise stress ECG | Peak voltage of STD during | 0.32 | 0.0046 |
|  | Stress ECG |  |  |
|  | ST/HR slope | −0.046 | 0.68 |
|  | Recovery index | 0.043 | 0.70 |
| Echocardiogram | IVSTd | −0.19 | 0.094 |
|  | LVPWTd | −0.24 | 0.031 |
|  | LVDd | −0.0005 | 1.00 |
|  | LVM | −0.26 | 0.018 |
| Note: p-values are obtained by comparison of the voltage of ST-segment depression on resting electrocardiogram and parameters of basic characteristics, resting and stress electrocardiogram and echocardiogram by regression analysis. Abbreviations: ECG, electrocardiogram; IVSTd, interventricular septal wall thickness at end-diastole; LVDd, left ventricular internal diameter at end-diastole; LVPWTd, left ventricular posterior wall thickness at end-diastole; LVM, left ventricular mass; STD, ST-segment depression; ST/HR slope, ST-segment/heart-rate slope |  |  |  |

### 3.5 Receiver operating characteristic curves of the different parameters on resting electrocardiogram for left ventricular mass on echocardiogram

Receiver operating characteristic (ROC) curves were then constructed for evaluating LVM on Echo to determine the highest area under curve (AUC) for different parameters on resting ECG (Figure 4). According to ROC curves, the best indicator for diagnosing LVH using Echo with LVM was the STD voltage (AUC = 0.62). In contrast, the Sokolow-Lyon’s voltage (AUC = 0.54, p = 0.45) and the Romhilt-Estes score (AUC = 0.5, p = 0.30) had lower diagnostic ability than the voltage of STD, however, the difference was not significant.

**Figure 4.**
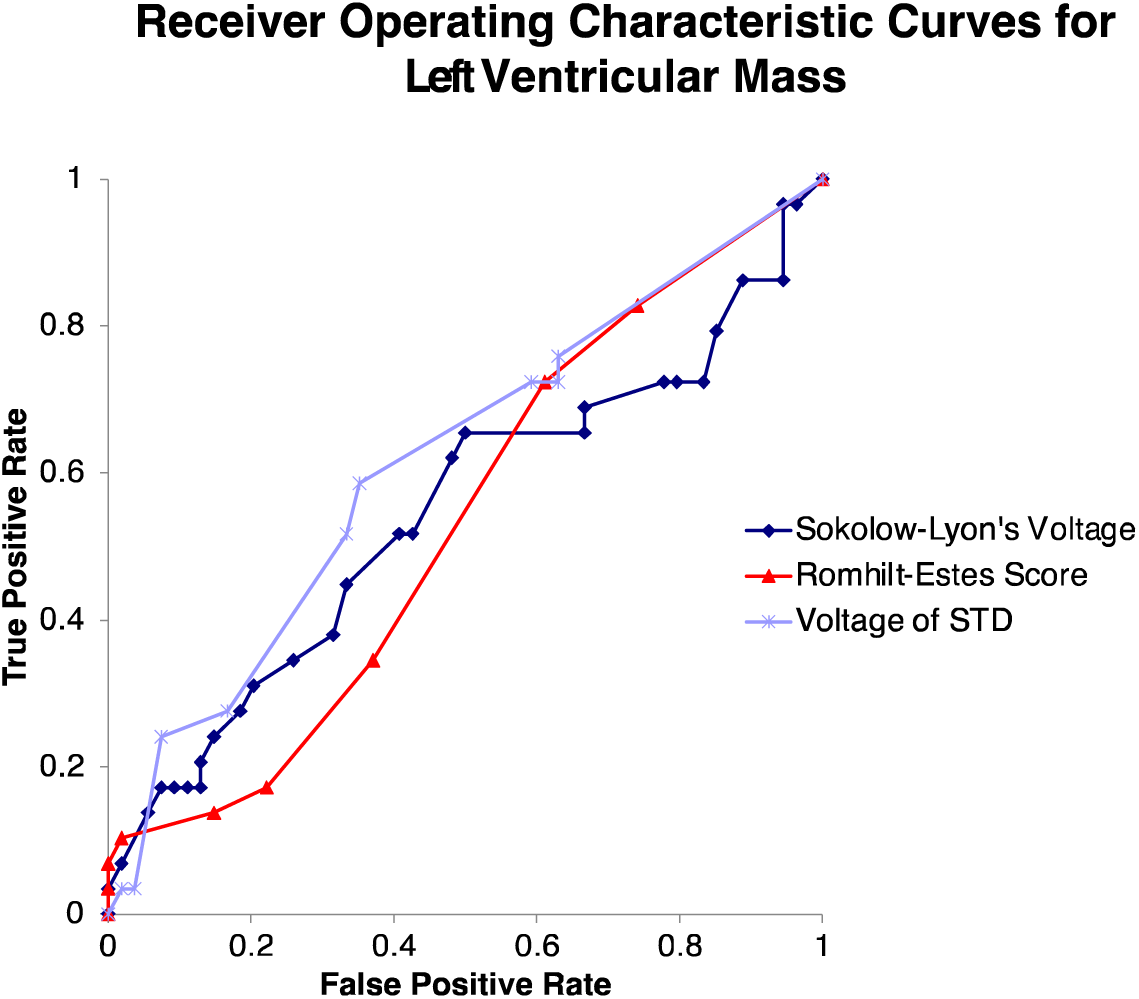
Receiver operating characteristic curves of electrocardiographic parameters — voltage of ST-segment depression, Sokolow-Lyon’s voltage, Romhilt-Estes score and echocardiographic parameter — left ventricular mass determined by Lang^9^

### 3.6 Multivariate logistic regression analysis with voltage of ST-segment depression

Multivariate analysis was performed to corroborate the independent statistically significant effects of the parameters. STD voltage was significantly correlated with age and peak STD voltage on stress ECG, but not with other parameters, using multivariate logistic regression analysis (Table 4). The likelihood ratio was 20.77 (p < 0.001).

**Table 4.**
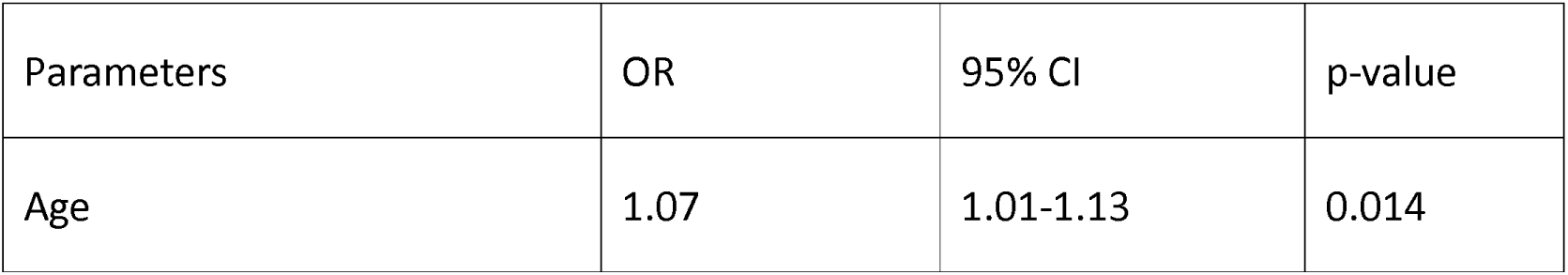
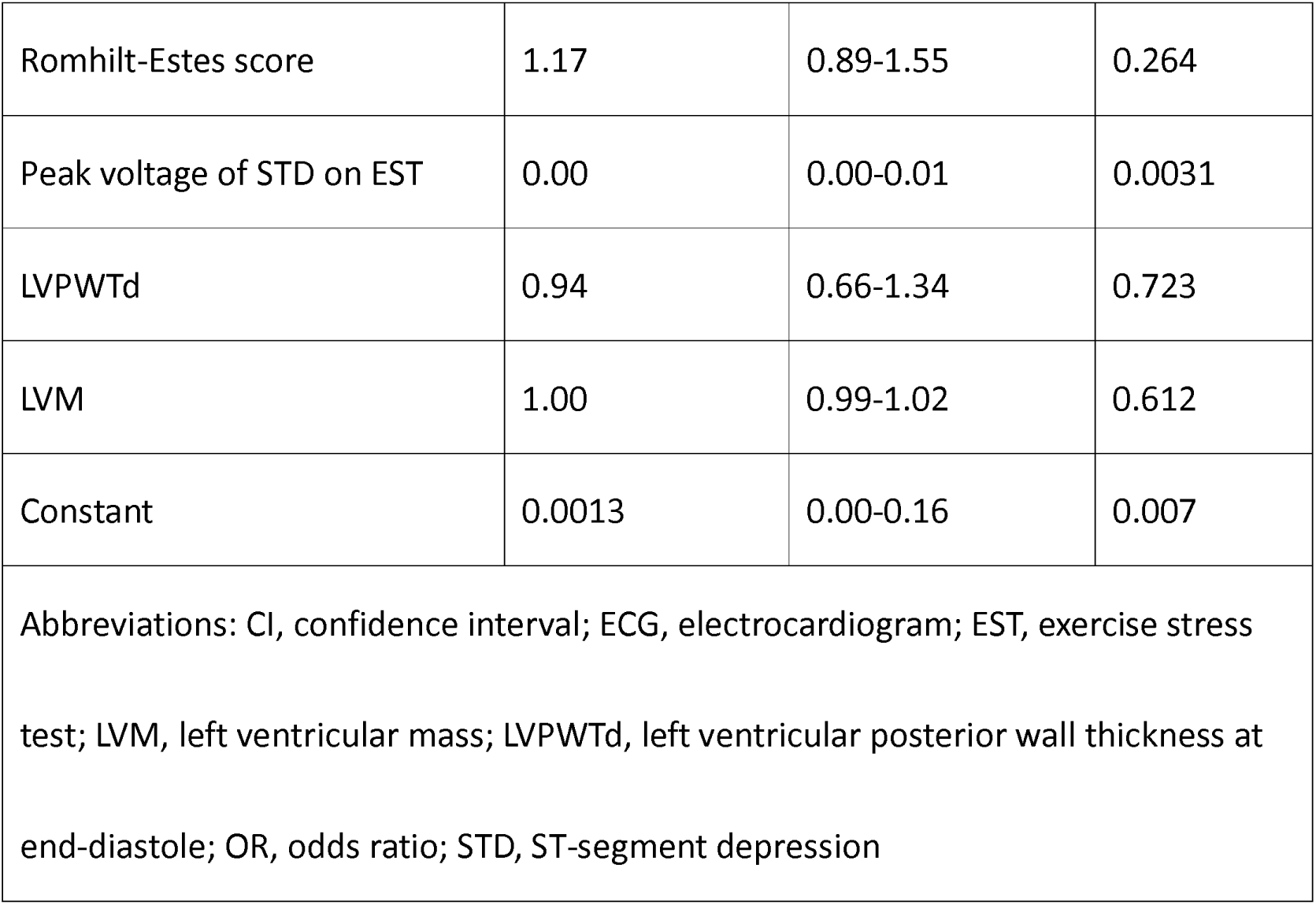
Multivariate logistic regression analysis of voltage of ST-segment depression on resting electrocardiogram with parameters of basic characteristics, resting and stress electrocardiogram and echocardiogram.

| Parameters | OR | 95% CI | p-value |
| --- | --- | --- | --- |
| Age | 1.07 | 1.01-1.13 | 0.014 |
| Romhilt-Estes score | 1.17 | 0.89-1.55 | 0.264 |
| Peak voltage of STD on EST | 0.00 | 0.00-0.01 | 0.0031 |
| LVPWTd | 0.94 | 0.66-1.34 | 0.723 |
| LVM | 1.00 | 0.99-1.02 | 0.612 |
| Constant | 0.0013 | 0.00-0.16 | 0.007 |
| Abbreviations: CI, confidence interval; ECG, electrocardiogram; EST, exercise stress test; LVM, left ventricular mass; LVPWTd, left ventricular posterior wall thickness at end-diastole; OR, odds ratio; STD, ST-segment depression |  |  |  |

### 3.7 Coronary arteriography

Six patients underwent coronary arteriography because of positive stress ECG results on EST. Coronary arteriography revealed CAD in two patients and normal coronary arteries in four patients (Table 2). Of the patients with CAD, one patient showed 75% stenosis in segment 7, and another showed 75% stenosis in segment 13^14^. Both patients with CAD reported non-anginal chest pain. None of the symptoms, risk factors of CAD and results, and parameters of stress ECG showed a significant correlation with results of coronary arteriography.

## 4 Discussion

This study examined patients with resting ECG, who reported no history of cardiac disease and exhibited non-anginal chest symptoms or were asymptomatic. Data was obtained by combining non-invasive cardiac screening methods, such as Echo and cycle ergometer stress ECG, with resting ECG. Among study patients, STD on resting ECG revealed an independent association with older age and, although not independently, LVH on ECG and Echo.

Combining non-invasive cardiac screening methods, such as Echo and cycle ergometer stress ECG, with resting ECG is a safe way to detect cardiac diseases and valuable for accurately diagnosing these conditions. Despite these clinical advantages, to the best of our knowledge, no previous study has assessed the utility of combining these three tests. Moreover, guidelines for Echo and stress ECG do not address combining the results of these tests for a more accurate ST-segment change evaluation^15, 16^. This is the first study to demonstrate that, on resting ECG, STD among patients with no history of cardiac disease or angina might imply LVH on Echo, by combining the results of resting and stress ECG and Echo. The findings of this study may be useful to investigate the cause of STD on resting ECG.

### 4.1 ST-segment depression on resting electrocardiogram in the literature

The cause of STD on resting ECG in patients with no history of cardiac disease or symptoms remains undetermined.

A study by Short assessed resting ECG data and reported that 20% of all patients with ST-segment changes on resting ECG had LVH; 60% of the patients with ST-segment changes had CAD^17^. These findings may indicate a cause of ST-segment changes on resting ECG. However, the diagnostic criteria for LVH were based on the ECG findings in the study above. In this study, an LVM on Echo was adopted as a standard diagnostic criterion as an accurate method of diagnosing LVH on Echo^18, 19^.

In a study by Short, CAD was diagnosed as an anginal attack^17^. In this study, stress ECG findings were examined to detect silent myocardial ischemia for the patients before the occurrence of coronary heart disease events. Furthermore, in this study, for a more precise diagnosis of CAD (silent myocardial ischemia) on stress ECG, a ST-segment/heart-rate loop method was adopted during and in the recovery phase of exercise^20, 21^. The ST-segment/heart-rate loop method during exercise detects the presence and severity of CAD^20^; this method can discriminate non-ischemic (false positive) and ischemic (true positive) ST-segment changes^21^ most accurately^11^, during and at the recovery period of exercise.

In clinical practice, further assessment by contemporary Echo and cycle ergometer stress ECG may be more valuable for prompt and accurate detection of LVH and/or CAD. For stress ECG, evaluation using the heart-rate-adjusted ST-segment loop method might be more useful for detecting silent myocardial ischemia.

### 4.2 Older age and electrocardiographic changes

In this study, STD voltage on resting ECG in patients without history of cardiovascular disease or anginal chest pain was suggested to be independently correlated with age. In a review which address ECG changes in elderly patients, non-specific ST-T wave changes were found in one-sixth of subjects over 70 years old; this was considered to be due to non-cardiac factors, such as obesity, metabolic disorders, and aging changes in the chest^22^. The prevalence of ST-junction and segment depression on resting ECG was observed to be significantly higher in older age, and mortality in men with ST-junctional depression was higher^23^. STD in elderly people might need careful follow-up.

### 4.3 Left ventricular hypertrophy and patient prognosis

In this study, although not independently, STD was significantly correlated with LVM on Echo. Therefore, STD on resting ECG suggests the presence of LVH: a significant risk factor for cardiovascular events^24, 25^. Early detection of LVH could help prevent adverse cardiovascular events. From the ROC curves, STD tended to correlate with LVH on Echo than other ECG parameters. Therefore, STD on resting ECG may lead to early detection of LVH, which may well be beneficial.

### 4.4 Utility of electrocardiogram in diagnosing left ventricular hypertrophy

Diagnosis of LVH on ECG has been primarily based on QRS voltages, such as Sokolow-Lyon’s voltage, Romhilt-Estes score, and Cornell voltage. Despite having higher specificities, these criteria consistently have low sensitivities^26^. Several ECG findings other than QRS voltage have been proposed to assess LVH, including strain-type ST-T abnormality^27^, left axis deviation^28^, and duration of QRS^29^. Resting ECG findings can be used to diagnose LVH, but only where valuable, specific, established criteria are used and the diagnostic values of these criteria are not compared^26^.

Diagnosing LVH using ECG may be challenging given that the sensitivity is low and no consensus has been reached regarding criteria^30^. Nevertheless, ECG continues to be a valuable modality for diagnosing LVH, as it is widely available, low cost, and can detect electrical changes secondary to LVH.

### 4.5 ST-segment depression for diagnosis of left ventricular hypertrophy using electrocardiogram

STD is related to LVH, with Gubner et al. reported the following ECG findings as diagnostic: STD in I lead, even with depth of 0.05 mV; T-wave inversion of more than 0.10 mV in V_1_ lead; voltages of the R wave on lead I and S wave on lead III > 2.2 mV, when accompanied by left axis deviation^28^.

Notably, they also reported that these ECG changes could be observed before emergence of LVH on ECG^28^. Regarding the Romhilt-Estes score, the presence of STD was included as a diagnostic criterion for LVH. In this study, STD suggested LVH on Echo. According to ROC curves, the AUC for STD was larger than that for Sokolow-Lyon’s voltage and Romhilt-Estes score. Therefore, STD voltage on resting ECG in patients without prior cardiac disease might be as accurate as other resting indices of LVH.

### 4.6 ST-segment depression on resting electrocardiogram and myocardial ischemia

In patients who underwent treadmill stress test for the diagnosis of chest pain, those with STD on resting ECG had CAD and, especially, three-vessel disease^31^. The results of the aforementioned study differ from those in this study. Patients in the previous study had chest pain suggestive of angina pectoris, which may explain the difference in results; the patients in this study had non-anginal pain or were asymptomatic.

In this study, cycle ergometer was adopted as a modality of stress ECG for nine tenths of the patients. Cycle ergometer has a limitation in a point that leg fatigue limits the exercise tolerance of the test, some of the patients in this study might have not reached their true limit of their exercise tolerance^16^.

In a review assessed the validity of stress ECG testing in patients with suspected CAD, it was useful for initial evaluation. On stress ECG, exercise-induced ST-segment changes, exercise capacity, and heart-rate response during the recovery period were obtained by stress ECG and useful for patient risk stratification^32^.

### 4.7 Heart-rate-adjusted analysis of ST-segment depression on stress ECG for detecting coronary artery disease

In this study, heart-rate-adjusted ST-segment depression analysis was used to interpret stress ECG testing, as this method can more accurately detect CAD^10^. Heart-rate-adjustment of exercise-induced ST-segment depression considers myocardial oxygen demand when deciding whether ST-segment depression on exercise testing is due to myocardial ischemia. Excessive ST-segment depression, divided by differences in heart rate between peak exercise and at rest, might be larger in patients with CAD^10^.

Furthermore, on stress ECG evaluation of CAD, heart-rate-adjusted ST-segment changes during both exercise and the recovery period were valuable in the detection of CAD^11^. Heart-rate-adjusted ST-segment depression is useful for discriminating ischemic and non-ischemic ST-changes during the exercise and the recovery periods^20^ and improves long-term prognostic values^33^.

### 4.8 ST-segment depression location

The location of STD on resting ECG can suggest whether the changes are related to CAD. Short reported that minor ST-T changes were most commonly located in the lateral leads in 81% of cases, either exclusively or in combination with inferior and/or anterior leads^17^. These findings are compatible with the results of this study, as the minor ST-segment and T-wave changes do not correspond to the respective coronary artery lesions. However, two-thirds of the patients in Short’s study had anginal chest pain and those who show junctional STD were excluded in that study. This might explain the difference in the prevalence of angina pectoris between Short’s study and this study.

### 4.9 Clinical implications

One important question posed by this study was how to deal with patients exhibiting minor asymptomatic ST-segment changes in routine clinical practice. Chou et al. reported that abnormalities in resting ECG were associated with an increased risk of subsequent cardiovascular events ^34^. However, they also stated that the clinical application of these findings was unclear. In this study, STD suggested the presence of LVH in asymptomatic patients and those with non-anginal chest pain without prior cardiac disease. Since the presence of LVH might indicate a risk of future cardiovascular events, patients with STD warrant careful follow-up, particularly those with risk factors such as hypertension and diabetes mellitus.

To accurately detect covert cardiac diseases, it is important to consider risk factors which can be addressed. Even if asymptomatic, resting ECG is recommended for patients with hypertension and/or diabetes mellitus^35^. Additionally, the presence of ST-segment changes in patients with hypertension and/or diabetes mellitus warrants further investigation, including Echo and stress ECG.

### 4.10 Limitations

The main limitation of this study was that the type of STD was not considered in patient enrollment. Classification of STD type is important when evaluating its significance. The Minnesota Code is widely used for differentiating ECG changes and is based on a strict criterion for the classification of ECG changes^36^. Nevertheless, in this study, STD voltages correlated with LVM on Echo, without considering the types of STD on resting ECG.

## 5 Conclusion

STD on resting ECG predominantly implied LVH on Echo, based patients without history of cardiac disease who experienced non-anginal chest pain or were asymptomatic. This is the first study to elucidate the cause of STD on ECG screening using a combination of three modalities: resting and stress ECG and Echo. STD on resting ECG also correlated with older age. These findings may be helpful for interpreting ST-segment changes in clinical settings.

## Data availability statement

Data supporting the findings of this study is available from the corresponding author, MM, upon request.

## Ethical Approval

This study is conducted according to Declaration of Helsinki and approved by ethical committee of Saito Hospital.

## Consent

This study is retrospective, observational study and written informed consent was not needed.

## Data Availability

All data produced in the present study are available upon reasonable request to the authors

## Nonstandard Abbreviations and Acronyms

AUC: Area under curve
CAD: Coronary artery disease
Echo: Echocardiography
LVH: Left ventricular hypertrophy
LVM: Left ventricular mass
LVPWTd: Left ventricular posterior wall thickness at end-diastole
STD: ST-segment depression
ST/HR slope: ST-segment/heart-rate slope

## Acknowledgment

None.

## Supplemental Files

None

